# SARS-CoV-2 IgM and IgG serology and clinical outcomes in COVID-19 patients

**DOI:** 10.1101/2022.08.21.22279044

**Authors:** S Mohan, Pratap Kumar, Prasan Kumar Panda, Vikram Jain, Rohit Raina, Sarama Saha, S Vivekandan, Balram J Omar

## Abstract

**Background:** The SARS-CoV-2 virus has become pandemic for the last 2 years. Inflammatory response to the virus leads to organ dysfunction and death. Predicting the severity of inflammatory response helps in managing critical patients using serology tests IgG and IgM. We conducted a longitudinal study to correlate serum SARS-CoV-2 IgM and IgG serology with clinical outcomes in COVID-19 patients.

**Methods:** We analyzed patient data from March to December of 2020 for those who were admitted at AIIMS Rishikesh. Clinical and laboratory data of these patients were collected from the e-hospital portal and analysed. Correlation was seen with clinical outcomes and was assessed using MS Excel 2010 and SPSS software.

**Results:** Out of 494 patients, the mean age of patients was 48.95 ± 16.40 years and there were more male patients in the study (66.0%). The patients were classified into 4 groups; mild-moderate 328 (67.1%), severe 131 (26.8%) and critical 30 (6.1%). The mean duration from symptom onset to serology testing was 19.87 ± 30.53 days. In-hospital mortality was observed in 25.1% patients. The seropositivity rate (i.e., either IgG or IgM >10 AU) was 50%. There was a significant difference between the 2 groups in terms of IgM Levels (AU/mL) (W = 33428.000, p = <0.001) and IgG Levels (AU/mL) (W = 39256.500, p = <0.001), with the median IgM/ IgG Levels (AU/mL) being highest in the RT-PCR-Positive group. There was no significant difference between the groups in terms of IgM Levels and IgG levels with all other clinical outcomes (disease severity, septic shock, Intensive care admission, mechanical ventilation and mortality).

**Conclusion:** Serology (IgM and IgG) levels are high in RTPCR positive group compared to clinical COVID-19. However, serology cannot be useful for the prediction of disease outcomes except few situations. The study also highlights the importance of doing serology at a particular time as antibody titres vary with the duration of the disease.

## Introduction

COVID-19 has affected almost 581 million people with around 6.4 million deaths as of July 2022 (WHO).(1) Severe acute respiratory syndrome coronavirus-2 (SARS-CoV-2) was identified as the causative agent. SARS-CoV-2 causes a respiratory infection with systemic involvement and an estimated 1% death rate. SARS-CoV-2 can infect individuals from different age groups and causes a wide spectrum of disease manifestations ranging from asymptomatic, mild, moderate to severe symptoms with possible fatal outcomes.(2) Age, sex, pre-existing comorbidities, host genetics as well as host immune response are the key factors determining the outcomes.(3)

The RTPCR assay is the right method to diagnose SARS-CoV-2. Unfortunately, the sensitivity of the RNA test in the real world is not satisfactory and, false-negative and false-positive cases have also been reported owing to several factors.(4) According to recent WHO case definitions, the RTPCR negative patients who meet clinical and epidemiological criteria or patients with severe acute respiratory illness who have typical chest imaging features or unexplained anosmia or ageusia are termed as probable COVID-19 patients.(5,6)

Serological tests are increasingly applied for the diagnosis of SARS-CoV-2 infection. Blood levels of immunoglobulin SARS-CoV-2 IgG & IgM, are also deployed for evaluating immune responses and confirming the diagnosis in symptomatic patients presenting outside of the window of positivity for RTPCR-based SARS-CoV-2 testing.(7) Few studies have assessed the utility of seroconversion profiles to predict infection severity or outcomes following SARS-CoV-2 infection. A strong association was observed between the magnitude of antibody response and patient survival, disease severity and fatal outcomes. Furthermore, several studies have documented discrepancy in findings related to the timing of SARS-CoV-2 antibody seroconversion and the onset of symptoms.(8-10) More information about the dynamics of the early humoral immune response is needed to realize the full potential of serological testing for SARS-CoV-2. The dynamics of antibody responses, in COVID-19 patients with different clinical presentations, is still not well-characterized. Such information can help our understanding of the nature of COVID-19 infection and guide patient management.

Here, we studied the seropositivity and kinetics of SARS-CoV-2 IgM and IgG antibodies in blood samples collected between 2 to 85 days post-symptoms onset from a cohort of 493 COVID-19 patients. The objectivity were correlation of the serology (IgM and IgG) with RTPCR status, disease severity (mild to critical), ICU admission, septic shock, acute kidney injury, and in hospital mortality.

## Material and methods

### Study design and setting

The study is a observational longitudinal study conducted on COVID-19 patients admitted in a tertiary care hospital, All India Institute of Medical Sciences (AIIMS), Rishikesh, India from August, 2020 to Nov, 2020. The study was designed according to the Strengthening the Reporting of Observational Studies in Epidemiology (STROBE) reporting guidelines.

### Inclusion criteria

1. COVID-19 patients with detectable SARS-CoV-2 RNA in respiratory samples since disease onset.
2. Clinical COVID-19 patients i.e. cases with clinical manifestations characteristic of COVID-19 but with negative SARS-CoV-2 RT-PCR test from admission until discharges (1,2).
3. Patients of both genders with age ≥15 years.

### Exclusion criteria

1. Patients not fulfilling COVID-19 diagnostic criteria as per institutional protocol.
2. Asymptomatic patients, pregnant women, and patients having incomplete data

### Case definitions

#### COVID-19 Severity classification

Patients were classified as mild, moderate, severe, and critical according to the WHO guidelines (1).

#### Serological tests

iFlash-SARS-CoV-2 (Shenzhen Yhlo Biotech Co. Ltd.), a paramagnetic particle based chemiluminescent immunoassay (CLIA) was used for the determination of IgM and IgG antibodies against SARS-CoV-2 nucleocapsid protein and spike protein. According to the manufacturer’s inserts (V1.0 English Fd. 2020–02-20), the IgM and IgG cut-off is 10AU/ml. i.e., an antibody titre above titre over 10AU/mL was regarded as positive.

#### Treatment of patients

Patients were treated uniformly as per institutional guideline.

#### Participants’ enrolment

All COVID-19 admitted patients at All India Institute of Medical Sciences, Rishikesh during above time period.

#### Variables and Outcome and Data collection

Full information regarding demographic characteristics, time course of symptoms, time of presentation and testing, presenting symptoms, final diagnosis, treatments received (i.e. oxygen therapy, corticosteroids, ICU admission, invasive ventilation requirement and dialysis) were collected in master excel. The medical records were further critically reviewed for important missed data.

#### Study size

All consecutive patients during above time period.

#### Ethics

The Approval for this study was obtained from institute ethics committee of AIIMS Rishikesh with approval no CTRI/2020/08/027169.

#### Statistical methods

All the statistical analyses were performed using statistical package for social sciences (SPSS), Windows version 23 software package (SPSS, CHICAGO, IL, USA). Non-normally distributed continuous variables were presented as medians (interquartile ranges [IQR]). Differences between non-normally distributed continuous variables were assessed using the Mann–Whitney U test. Categorical variables were presented as counts (%). Differences between categorical variables were assessed using the χ^2^ or Fisher’s exact tests. A two-sided value of *p*⁏<⁏0.05 was considered statistically significant.

#### Bias

As all patients sampling for IgG and IgM was conducted only once, and time to sampling may be an important variable which can confound the study results, we analyzed the association between different clinical outcomes and its association with IgG and IgM levels in a time dependent manner on the basis of time interval between symptom onset and IgM and IgG testing. As there is no gold standard diagnostic test for diagnosis of COVID-19, we used Bayesian latent class modelling for evaluation of the diagnostic performance of RT-PCR, IgM and IgG test in COVID-19.

## Results

### Demographic characteristics

A total of 494 hospitalized patients were enrolled in the retrospective study, among them 199 were RTPCR positive and 294 were clinically diagnosed COVID-19 patients (Table 1) (Figure 1 and 2).

**Table 1:** Basic characteristics of the cohort.

|  |  |
| --- | --- |
| Features | Mean ± SD \|\| Median (IQR) \|\| Min-Max \|\| Frequency (%) |
| Age (Yrs) | 48.95 ± 16.40 \|\| 50.00 (36.00-61.00) \|\| 18.00 - 87.00 |
| **Gender** |  |
| Male | 326 (66.0%) |
| Female | 168 (34.0%) |
| **IgG(AU/mL)** | 29.66 ± 37.09 \|\| 7.82 (0.63-57.07) \|\|<br>0.00 - 176.60 |
| **IgG** |  |
| <10 AU/mL | 257 (52.0%) |
| >10 AU/mL | 237 (48.0%) |
| **IgM (AU/mL)** | 30.14 ± 109.31 \|\| 0.96 (0.48-7.68) \|\|<br>0.16 - 949.66 |
| **IgM** |  |
| <10 AU/mL | 358 (77.2%) |
| >10 AU/mL | 106 (22.8%) |
| **RT-PCR** |  |
| Positive | 199 (40.4%) |
| Negative | 294 (59.6%) |
| **Onset-Testing Interval (Days)** | 19.87 ± 30.53 \|\| 12.00 (7.00-21.00) \|\|<br>1.00 - 302.00 |

**Figure 1:**
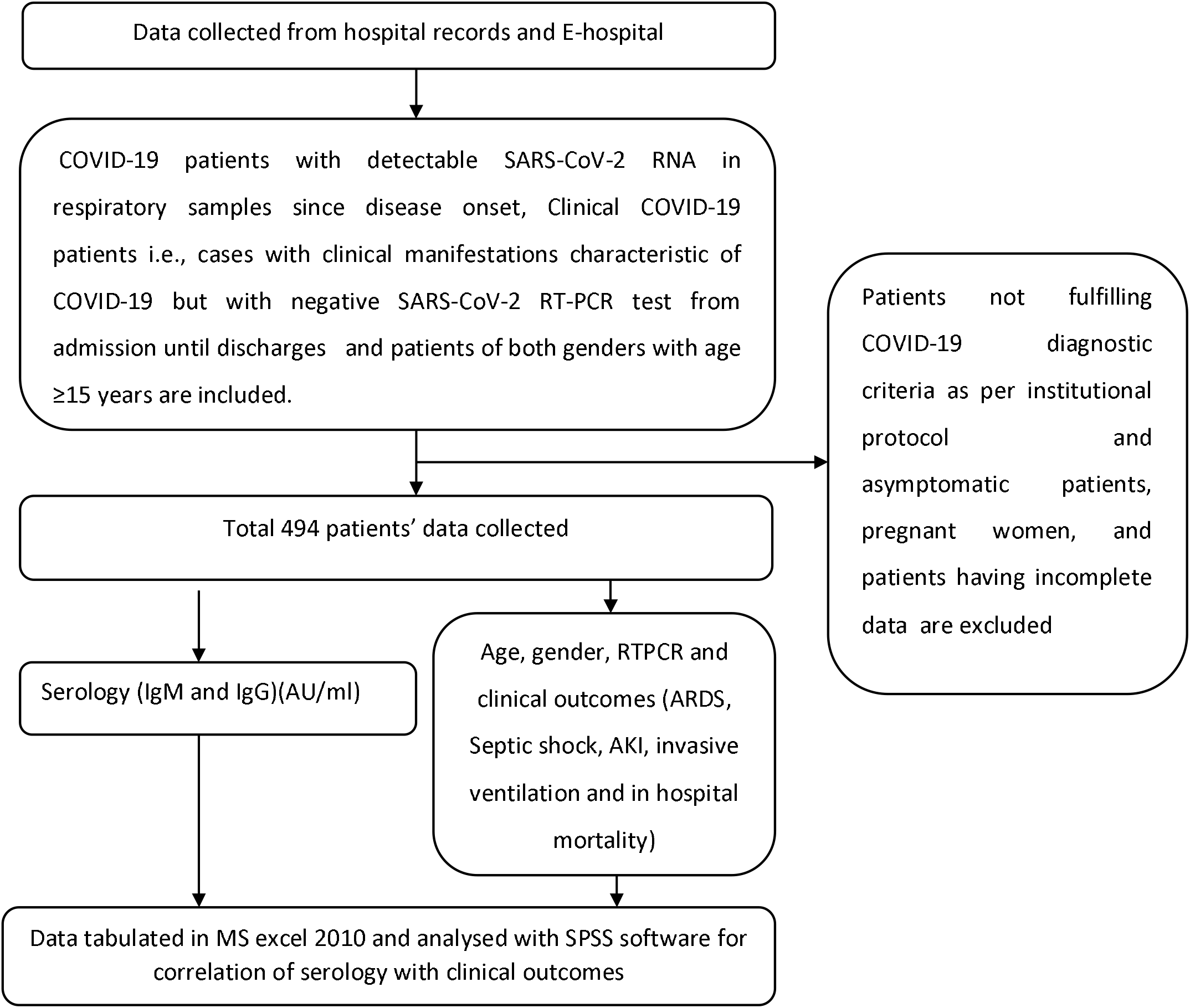

**Figure 2:**
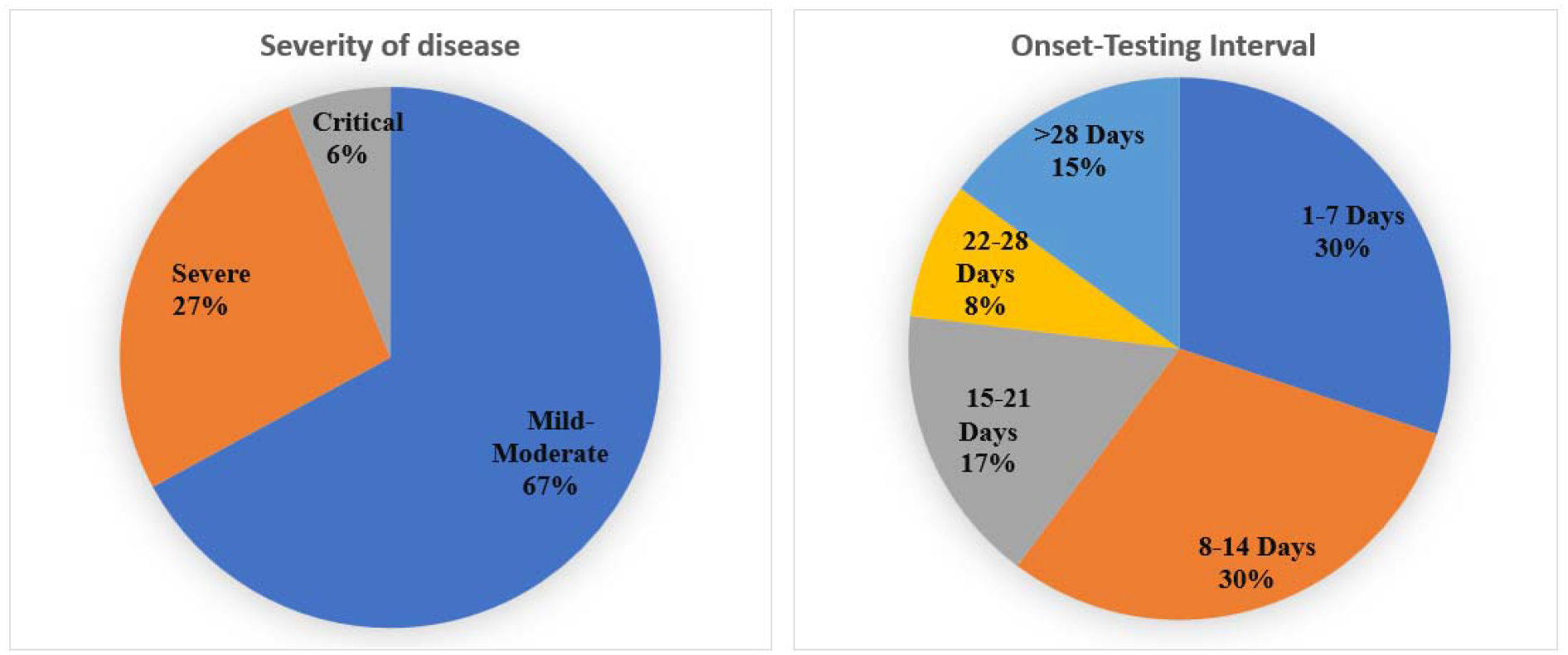

### Seropositivity status among COVID-19 patients

In this cohort of 494 patients, the seropositivity rate (i.e. either IgM or IgG >10 AU) was 247 (50%). Out of these IgM seropositivity was observed in 106/494 (47.97%) and for IgG 237/494 (21.45%). The Seropositivity rate was more for IgG compared to IgM. IgM or IgG seropositivity increased to a peak at week 4 then decreases after 4 weeks (> 28 days) (Figure 3) (Table 2 & 3).

**Figure 3:**
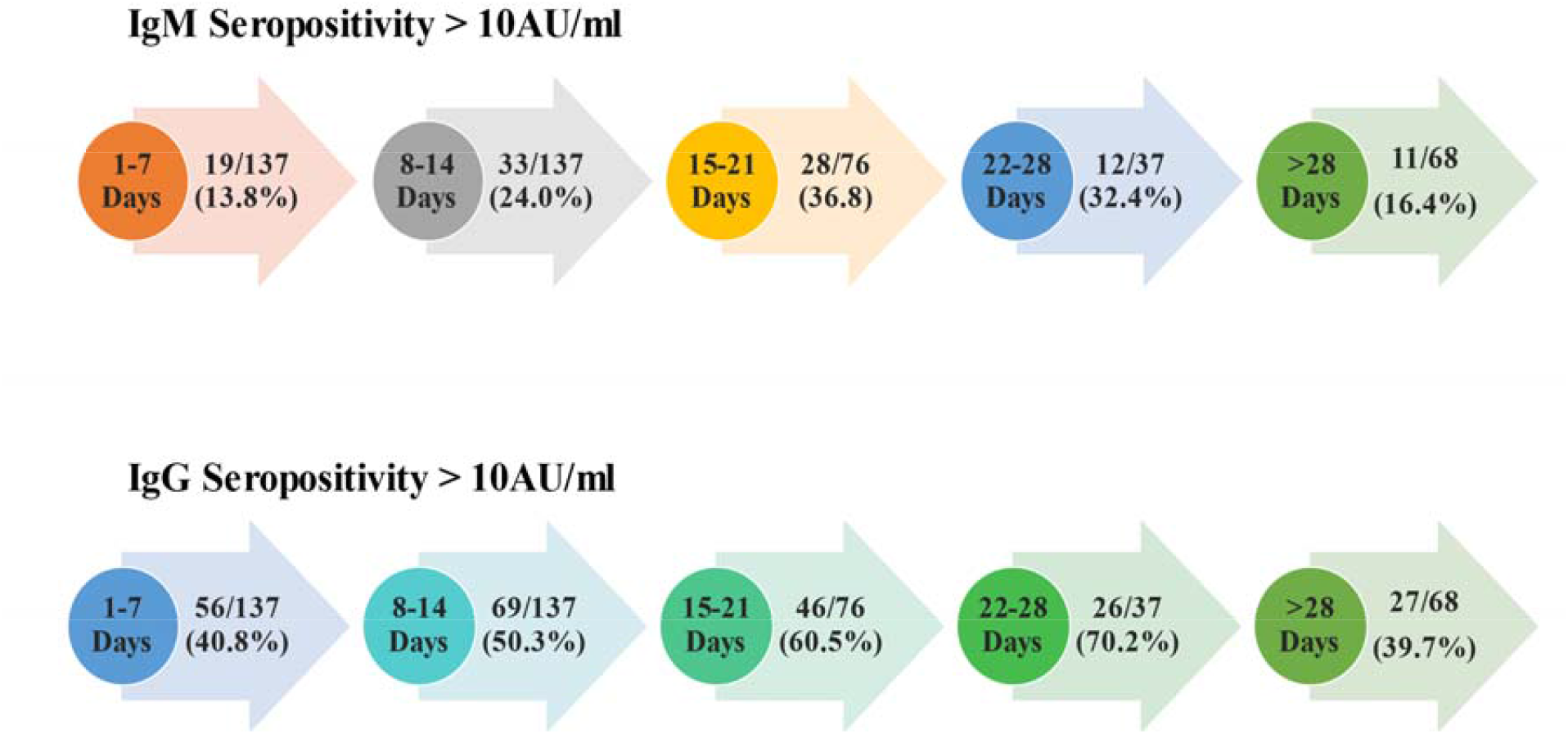

**Table 2:**
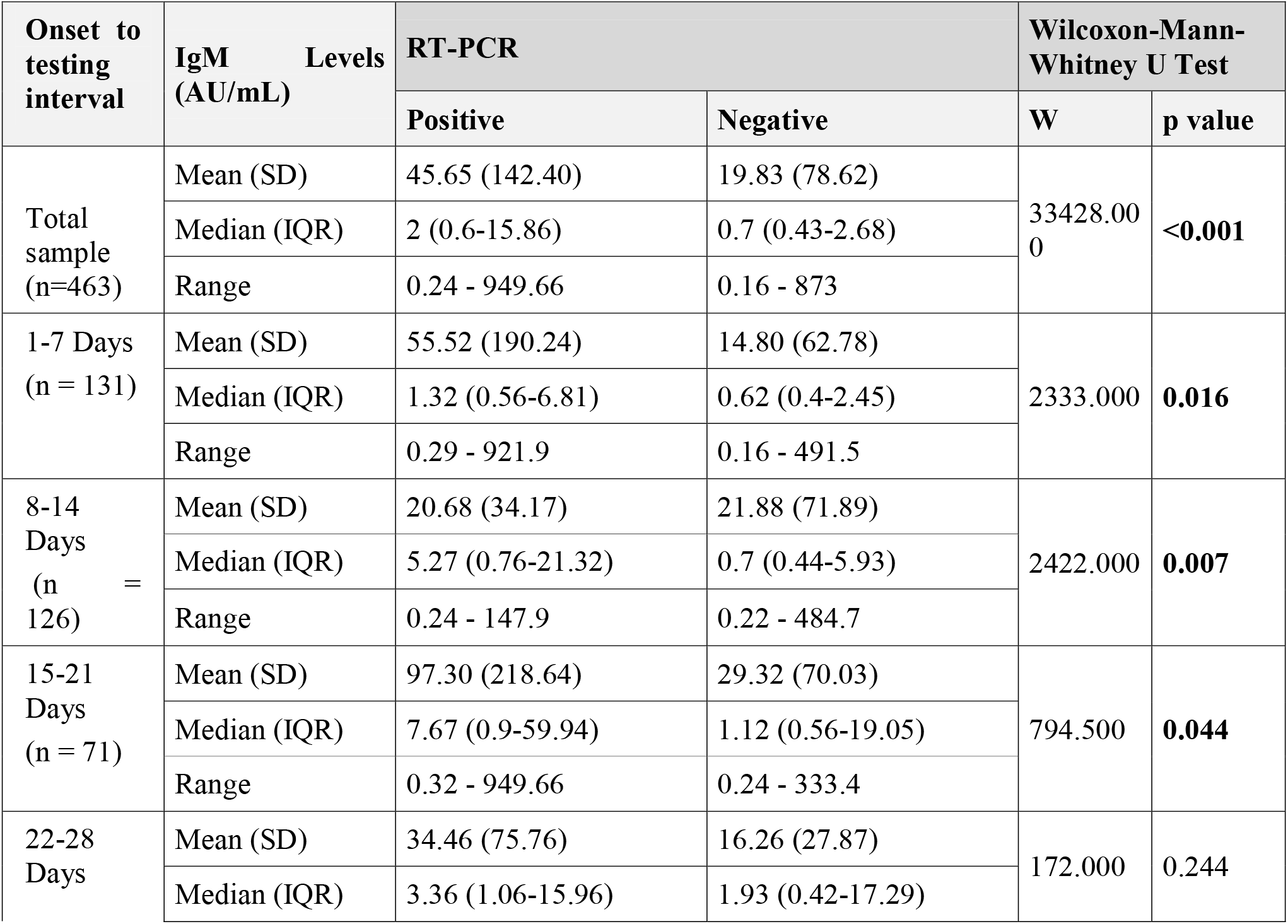
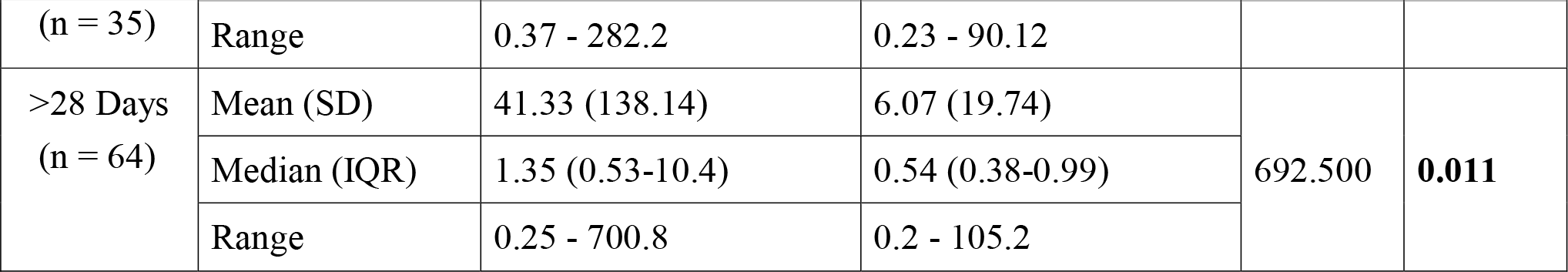
Comparison of the 2 Subgroups of the Variable RT-PCR in Terms of IgM Levels (AU/mL) :

**Table 3:**
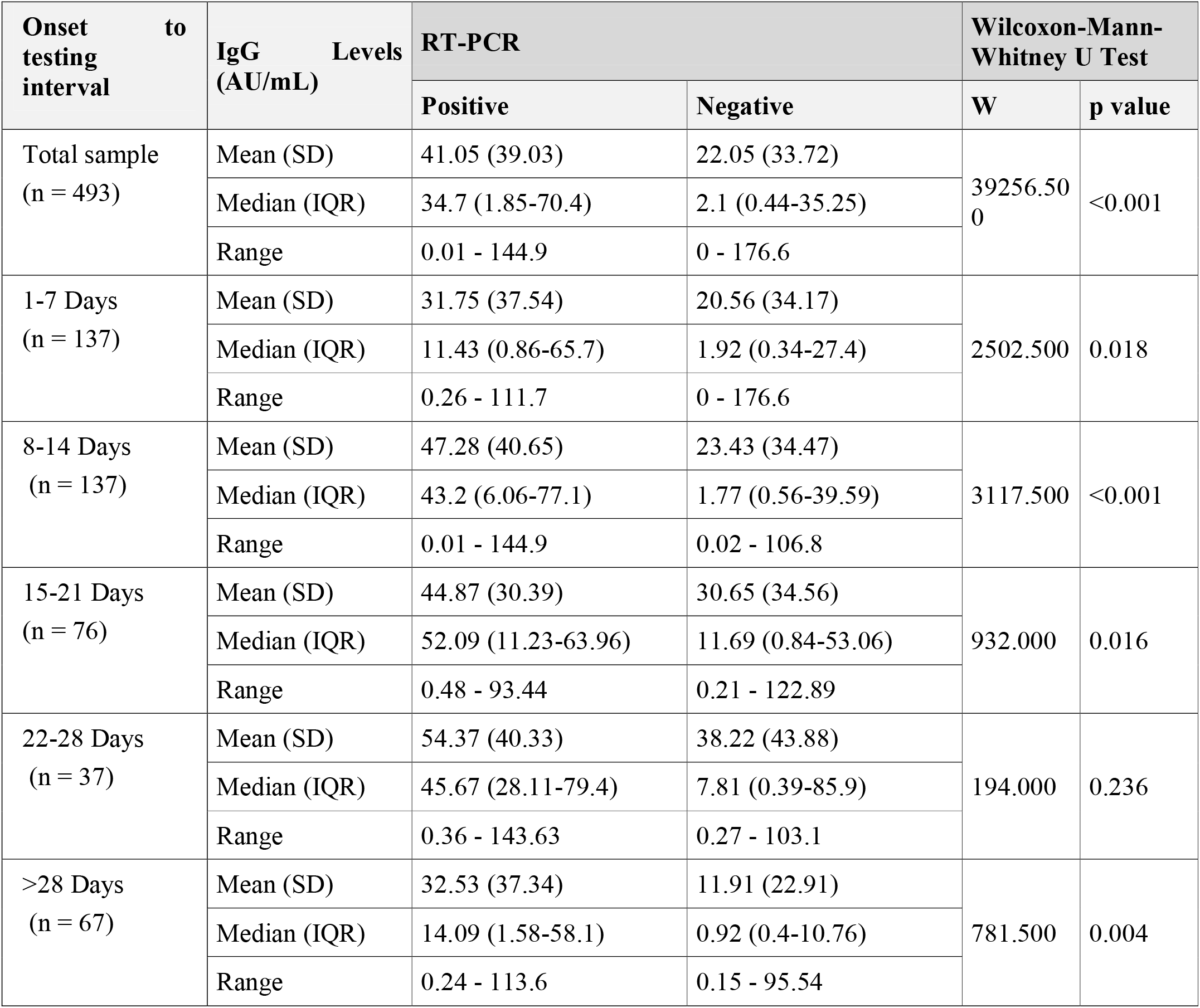
Comparison of the 2 Subgroups of the Variable RT-PCR in Terms of IgG Levels (AU/mL)

### Association between COVID-19 serology and RTPCR status

There was a significant difference between the 2 groups in terms of IgM Levels (AU/mL) (W = 33428.000, p = <0.001) and IgG Levels (AU/mL) (W = 39256.500, p = <0.001), with the median IgM/ IgG Levels (AU/mL) being highest in the RT-PCR-Positive group.

In all weeks There was a significant difference between the 2 groups in terms of IgM and IgG Levels (AU/mL) with the median IgM/ IgG Levels (AU/mL) being highest in the RT-PCR-Positive group. Except for week 4 (22-28 Days) there was no significant difference in terms of IgM and IgG Levels (AU/mL)

### Association between COVID-19 serology and disease severity

There was no significant difference between the groups in terms of IgM Levels (AU/mL) (χ2 = 2.975, p = 0.395) and IgG levels (χ2 = 2.463, p = 0.482).

In week 3, there was a significant difference between the 3 groups in terms of IgM Levels (AU/mL) (χ2 = 7.732, p = 0.021) and IgG Levels (AU/mL) (χ2 = 7.707, p = 0.021), with the median IgM and IgG Levels (AU/mL) being highest in the critical group. In all the other weeks There was a significant difference between the 2 groups in terms of IgM and IgG Levels (AU/mL)

### Association of COVID-19 serology with ARDS types and Oxygen requirement

There was a significant difference between the 4 groups in terms of IgM Levels (AU/mL) (χ2 = 7.985, p = 0.046) and IgG Levels (AU/mL) (χ2 = 8.501, p = 0.037). The median IgM Levels (AU/mL) being highest in the Mild ARDS group and median IgG Levels (AU/mL) being highest in the Moderate ARDS group.

In all weeks no significant difference between the groups in terms of IgM levels and IgG levels. However, in week 3 there was a significant difference between the 4 groups in terms of IgM Levels (AU/mL) (χ2 = 10.837, p = 0.013) and IgG of IgG Levels (AU/mL) (χ2 = 9.682, p = 0.021). The median IgM Levels (AU/mL) being highest in the Mild ARDS group and the median IgG Levels (AU/mL) being highest in the severe ARDS group.

There was a significant difference between the 3 groups in terms of IgM Levels (AU/mL) (χ2 = 6.795, p = 0.033), with the median IgM Levels (AU/mL) being highest in the Oxygen Therapy: <6 L/min group. There was no significant difference between the groups in terms of IgG Levels (AU/mL) (χ2 = 4.532, p = 0.104).

There was a significant difference between the 3 groups in terms of IgM Levels (AU/mL) in week 1 (χ2 = 6.053, p = 0.048), with the median IgM Levels (AU/mL) being highest in the Oxygen Therapy: <6 L/min group, week 2 (χ2 = 6.392, p = 0.041), with the median IgM Levels (AU/mL) being highest in the Oxygen Therapy: >6 L/min group and Week3 (χ2 = 6.283, p = 0.043), with the median IgM Levels (AU/mL) being highest in the Oxygen Therapy: <6 L/min group. There was a significant difference between the 3 groups in terms of IgG Levels (AU/mL) (χ2 = 8.629, p = 0.013), with the median IgG Levels (AU/mL) being highest in the Oxygen Therapy: >6 L/min group. In all other weeks no significant difference between the groups in terms of IgM levels and IgG levels

### Association of COVID-19 serology with Septic shock

There was no significant difference between the groups in terms of IgM Levels (AU/mL) (W = 1191.500, p = 0.168) and IgG Levels (AU/mL) (W = 19537.500, p = 0.261).

In all weeks no significant difference between the groups in terms of IgM levels and IgG levels. However, there was a significant difference between the 2 groups in terms of IgM Levels AU/mL) (W = 1827.000, p = 0.035), with the median IgM Levels (AU/mL) being highest in the no Septic Shock group. In week 3 IgG Levels (AU/mL) (W = 317.000, p = 0.022), with the median IgG Levels (AU/mL) being highest in the Septic Shock group and in > 4 week (W = 366.000, p = 0.042), with the median IgG Levels (AU/mL) being highest in the no Septic Shock group.

### Association of COVID-19 serology with requirement of ICU admission

There was no significant difference between the groups in terms of IgM Levels (AU/mL) (W = 23685.000, p = 0.668) and IgG (W = 25763.500, p = 0.157).

In all weeks no significant difference between the groups in terms of IgM levels and IgG levels. However, there was a significant difference between the 2 groups in terms of IgM Levels (AU/mL) on week 3 (W = 403.500, p = 0.031) and IgG) (W = 460.000, p = 0.038) with the median IgM Levels (AU/mL) being highest in the group requiring ICU admission.

### Association of COVID-19 serology with requirement of mechanical ventilation

There was no significant difference between the groups in terms of IgM Levels (AU/mL) (W = 20744.500, p = 0.099) and IgG Levels (AU/mL) (W = 23067.000, p = 0.460).

In all weeks no significant difference between the groups in terms of IgM levels and IgG levels. However, there was a significant difference between the 2 groups in terms of IgM Levels (AU/mL) on week 2 (W = 2070.000, p = 0.035) and > 4 week (>28days) (W = 358.500, p = 0.033), with the median IgM Levels (AU/mL) being highest in the no Invasive Ventilation group.

### Association of COVID-19 serology with acute kidney injury (AKI) and requirement of dialysis

There was no significant difference between the groups in terms of IgM Levels (AU/mL) (W = 23261.500, p = 0.425) and IgG Levels (AU/mL) (W = 26023.500, p = 0.767).

In all weeks no significant difference between the groups in terms of IgM levels and IgG levels. However, there was a significant difference between the 2 groups in terms of IgM Levels (AU/mL) on week 2 (W = 2473.000, p = 0.008), and IgG Levels (AU/mL) (W = 2755.500, p = 0.043) with the median IgM/ IgG Levels (AU/mL) being highest in the no Acute Kidney Injury group.

There was a significant difference between the 2 groups in terms of IgM Levels (AU/mL) (W = 14962.000, p = <0.001), with the median IgM Levels (AU/mL) being highest in the no Dialysis group. However, there was no significant difference between the groups in terms of IgG Levels (AU/mL) (W = 14553.000, p = 0.206). In all weeks no significant difference between the groups in terms of IgM levels and IgG levels.

### Association between COVID-19 serology and outcome: Survivor versus non-survivor

There was no significant difference between the groups in terms of IgM levels (AU/mL) (W = 21870.000, p = 0.058) and IgG levels (AU/mL) (W = 23088.500, p = 0.738).

In all the weeks there was no significant difference between the groups in terms of IgM levels and IgG levels. However, there was a significant difference between the 2 groups in terms of IgM Levels (AU/mL) on week 4 (W = 136.500, p = 0.032) and > 4 weeks (> 28days) (W = 575.500, p = 0.003) with the median IgM Levels (AU/mL) being highest in the survival group.

## Discussion

The RT-PCR test is the most commonly used molecular test for the diagnosis of COVID-19 infection and is considered the gold standard test.(11) COVID-19 serology has emerged as one of the alternatives for diagnosing the COVID disease. One of the meta-analyses by Chen m et al showed that the panel of IgG+ or IgM+ had a sensitivity of almost 79%, followed by IgG+ IgM+/-(73%), IgG+/-IgM+ (68%). Pooled specificities of these tests ranged from 98% to 100%.(12) In this study also, in patients who had clinical COVID-19, almost 50% of patients were seropositive (IgM+ or IgG+).

COVID-19 IgG can be used as a tool to predict the disease severity. One of the retrospective studies done by yan x et al, showed that patients who had severe COVID-19 disease had higher COVID-19 IgG antibodies after 1 year.(13) In this study also patients who were RT-PCR positive had statistically significant COVID-19 antibody serology. Also, Seropositivity for IgG increases as disease severity increases as shown in this study.

In one of the cross-sectional studies done in Iran, the study suggested that the patients who were IgG and IgM positive had more severe symptoms compared to patients who had negative serology.(14) If we see the relationship between the COVID-19 serology and complications, not many studies had been done in the past. This study had shown that patients who had higher COVID-19 IgG levels at three weeks had more severe ARDS and oxygen requirements compared to other patients. We also observed that there was a statistically significant difference in IgG antibody titres between present or absence of septic shock at three weeks. A similar trend was seen for ICU admissions and the need for mechanical ventilation. Also, in patients, who developed AKI there was more IgG seropositivity than IgM.

Previous studies by Liu X. et al., 2020, Hou et al., 2020 and Zhang B. et al. (2020) showed that higher antibodies (IgM and IgG) levels are seen in patients with severe and critical patients compared to mild-moderate patients.(15-17) Chen, Hao, et al. 2021 study shows similar results as the above studies. However, the study showed antibody titres levels may vary and higher antibody titres were present in some patients’ mild-moderate patients than in severe and critical patients. These findings are due to variation in serology to symptom onset interval.(18) The study also did not find a statistically significant correlation between antibody tires with AKI, mechanical ventilation, ICU requirement, septic shock and mortality.

This study shows that higher body titres are associated with poor outcomes at a particular time serology to symptom onset interval. There are some limitations in this study first, its retrospective study and dynamic observation variation in antibody tires with the outcomes studied in a single patient. Second, there are limited patients in severe and critical patients compared to mild and moderate which may lead to biases in the results.

## Conclusion

Serology (IgM and IgG) are inflammatory markers of COVID-19. In this study showed that serology levels are high in RTPCR positive group compared to clinical COVID-19. However, serology cannot be useful for the prediction of disease outcomes. The study also highlights the importance of doing serology at a particular time as antibody titres vary with the duration of the disease. In week interval there were significant correlation with clinical outcomes and serology on week 3.

## Data Availability

It will be made available to others as required upon requesting the corresponding author.

## Contributors

All contributed to the data collection, data analysis, manuscript writing, critically reviewed the draft, and approved it for publication.

## Data sharing

It will be made available to others as required upon requesting the corresponding author.

## Acknowledgment

COVID care team was collecting data, special thanks to all of them.

